# Forecasting influenza incidence as an ordinal variable using machine learning

**DOI:** 10.1101/2023.02.09.23285705

**Authors:** Haowei Wang, Kin On Kwok, Steven Riley

**Affiliations:** School of Public Health, Imperial College London, UK; MRC Centre for Global Infectious Disease Analysis and Abdul Latif Jameel Institute for Disease and Emergency Analytics, Imperial College London, UK; JC School of Public Health and Primary Care, The Chinese University of Hong Kong, Hong Kong Special administrative regions, China; Stanley Ho Centre for Emerging Infectious Diseases, The Chinese University of Hong Kong, Hong Kong Special Administrative Region, China; Hong Kong Institute of Asia-Pacific Studies, The Chinese University of Hong Kong, Hong Kong Special Administrative Region, China

**Author notes:** Corresponding authors: Steven Riley, School of Public Health, Imperial College London, Norfolk Place, London, W2 1PG.

## Abstract

Many mechanisms contribute to the variation in the incidence of influenza disease, such as strain evolution, the waning of immunity and changes in social mixing. Although machine learning methods have been developed for forecasting, these methods are used less commonly in influenza forecasts than statistical and mechanistic models. In this study, we applied a relatively new machine learning method, Extreme Gradient Boosting (XGBoost), to ordinal country-level influenza disease data. We developed a machine learning forecasting framework by adopting the XGBoost algorithm and training it with surveillance data for over 30 countries between 2010 and 2018 from the World Health Organisation’s FluID platform. We then used the model to predict incidence 1- to 4-week ahead. We evaluated the performance of XGBoost forecast models by comparing them with a null model and a historical average model using mean-zero error (MZE) and macro-averaged mean absolute error (mMAE). The XGBoost models were consistently more accurate than the null and historical models for all forecast time horizons. For 1-week ahead predictions across test sets, the mMAE of the XGBoost model with an extending training window was reduced by 78% on average compared to the null model. Although the mMAE increased with longer prediction horizons, XGBoost models showed a 62% reduction in mMAE compared to the null model for 4-week ahead predictions. Our results highlight the potential utility of machine learning methods in forecasting infectious disease incidence when that incidence is defined as an ordinal variable. In particular, the XGBoost model can be easily extended to include more features, thus capturing complex patterns and improving forecast accuracy. Given that many natural extreme phenomena, such as floods and earthquakes, are often described on an ordinal scale when informing planning and response, these results motivate further investigation of using similar scales for communicating risk from infectious diseases.

**Author Summary:** Accurate and timely influenza forecasting is essential to help policymakers improve influenza preparedness and responses to potential outbreaks and allocate medical resources effectively. Here, we present a machine learning framework based on Extreme Gradient Boosting (XBoost) for forecast influenza activity. We used publicly available weekly influenza-like illness (ILI) incidence data in 32 countries. The predictive performance of the machine learning framework was evaluated using several accuracy metrics and compared with baseline models. XGBoost model was shown to be the most accurate prediction approach, and its accuracy remained stable with increasing prediction time horizons. Our results suggest that the machine learning framework for forecasting ILI has the potential to be adopted as a valuable public health tool globally in the future.

## Introduction

Influenza forecasting plays a critical role in helping healthcare planners and policymakers to improve response to large seasonal epidemics and to mitigate their impact in terms of morbidity and mortality as well as social and economic impacts. Before the COVID-19 pandemic, a contagious respiratory illness caused by influenza viruses posed continual threats globally, causing an estimated 1 billion cases and up to 650000 deaths annually [1,2]. Accurate and timely forecasting of influenza epidemics in terms of the start of the epidemic, the time and size of the peak, and the duration of the epidemic enable policymakers to take effective interventions and optimise the allocation of healthcare resources. For example, to conduct the maximum number of elective surgical procedures prior to opening up space in intensive care wards for community-acquired pneumonia patients. However, reliable influenza forecasts remain a substantial challenge due to the variation of dominant influenza strains and environmental factors affecting the outbreak intensity. [3–5].

Different analytical methods have been used as the basis for forecast models of influenza disease. These methods can be classified into two broad categories: mechanistic models and statistical methods [5–7]. Mechanistic models attempt to reproduce key features of the underlying mechanism of transmission. Typical examples include classic compartmental models [8–11] and agent-based models (ABMs) [12–15]. Statistical approaches are phenomenological and do not attempt to reproduce the transmission mechanism, such as autoregressive integrated moving average (ARIMA) [16,17] and Gaussian Process Regression (GPR) [18,19].

Machine learning (ML) models have gained much attention in recent years and have gradually become the third category of models. Given that machine learning is sometimes differentiated from traditional statistics for its focus on prediction, it is not surprising that these methods are being applied to forecasting influenza. Traditional statistical models have a longstanding emphasis on inference, which is achieved through creating and fitting project-specific probability models. They are often less well-suited to data with large sample sizes and input variables [20]. By contrast, ML methods are data-driven and avoid making prior assumptions about underlying correlations and rather employ algorithms to identify patterns in the data [21,22]. In addition, ML methods are flexible in taking different types of input variables and a huge number of observations into consideration to improve predictive performance. The usage of some popular machine learning and deep learning methods in influenza forecasting has been discussed, such as Long Short Term Memory (LSTM) [23], Support Vector Machine (SVM) [24–27], and neural networks [28–31]. These methods show consistent and high forecasting accuracy but also suffer from the risk of over-fitting. Many natural extreme phenomena, such as floods and earthquakes, are often described on an ordinal scale, but infectious disease incidence is usually described as a count or a percentage of a population. The use of the moving epidemic method is a notable exception, which is now routinely used to compare and communicate the current state of epidemics across nearby connected populations [32]. However, influenza forecasting has remained focused on non-ordinal target observations [33], limiting the range of analytical methods that can be applied.

Extreme Gradient Boosting (XGboost) is a decision-tree-based ensemble machine learning method employing the gradient boosting algorithm [34]. It has demonstrated good prediction performance on a wide range of problems in different industries, including finance, physics and clinical research (e.g. patient diagnosis) [35–38].

In this paper, we aim to construct an XGBoost model to make short-term predictions of the weekly influenza incidence as an ordinal variable, ranging from 1-to 4-week ahead of the current time. We also evaluate the performance of XGBoost by accuracy metrics and compare it with baseline models.

## Methods

### Data

We used a global web-based collection tool for epidemiological indicators and data on influenza, Flu Informed Decisions (FluID) dataset from the World Health Organization (WHO) [39]. This platform collects weekly Influenza-Like Illness (ILI) incidence data from WHO member countries and regions, which is either submitted on a weekly basis or obtained by the WHO from regional networks such as EUROFlu [2].

The FluID data was available from ISO year 2010 week 1 to 2017 week 52 and included data from 146 countries initially. To ensure sufficient data for model training, countries with less than 50% of the data were excluded. Further countries that did not have at least 10 weeks of data for each year between 2010 and 2017 were also excluded. After applying these criteria were applied to the dataset, 32 countries remained, primarily in Europe and North America.

The key field in the data was ILI incidence, which was an integer ranging from 0 to 44965 (in week 52 of 2017 in the USA). This field was transformed into an ordinal variable by discretizing into N different bins with equal intervals, using N = 10 as the default. The highest incidence data for each country was used to add 10% as the upper boundary. The range from 0 to the upper limit was then divided into ten equal ordinal intervals, each of which was mapped to an ordinal value from 1 to 10, 1 represented the lowest incidence level, while 10 represented the highest level. Note that each country’s classification was based on its own influenza incidence level, so the same category in two different countries corresponded to different absolute levels of incidence.

### Models

We used XGBoost to establish a model to predict short-term weekly influenza incidence levels [34]. XGBoost is an implementation of the gradient-boosted decision trees and has been developed to improve computation speed and predictive performance for a variety of problems, including classification and regression [40]. Gradient boosting is an algorithm where new models are added in an adaptive way based on the residuals or errors of predictions from prior models and then combined to make the final prediction. Boosting is an ensemble method that creates a strong prediction model by iteratively combining a number of weak classifiers. New weak classifiers are added to correct the errors made by existing models, and every new model is added sequentially until no further improvements can be achieved or until a maximum number of models is added. Gradient boosting uses a gradient descent algorithm to minimize the loss when adding new models, i.e., at every optimization step, only models that reduce the residual or errors are added. XGBoost uses second-order Taylor expansion to minimize the loss function and added regularization terms to prevent overfitting.

To further validate the effectiveness of XGBoost for influenza forecasting, we constructed two additional baseline models for comparison: 1) null model, in which the prediction of the target week is the same as the most recent available observation week, and 2) historical average model in which the prediction of the target week is the average of observations of the same weeks in other years. The usual definition for historical average models is to mean value over prior observations for that week of the year [41]. Here, we use a slightly different categorical definition of the incidence level with the highest frequency across the same week in the other years in the study. The aim of the historical average model is similar to that of the null model, which is to provide a baseline for comparison with the accuracy of the XGBoost model, reflecting how users might informally forecast the incidence levels for the next 1 to 4 weeks.

### Forecast and features

Our target was to predict ordinal categories for each week at the country level. We first did regular machine learning forecasting, in which we trained the model with a training set of fixed durations and then assessed the model’s performance on a testing set. We designed our analysis to use 5 years of training data to predict one year of outcomes. The first training set used data from 2010 to 2014, the second training set used data from 2011 to 2015, and so on (S1 Table).

For predictors in the short-term forecasting model we used: categorical incidence levels of the prior *n* week and prior (*n* + 1) weeks, the month of the year (from January to December) and the season (spring, summer, autumn, and winter), where *n* represents the n-week ahead forecast. In a 1-week ahead forecast, the model uses the incidence levels of the prior 1 week and the prior 2 weeks; similarly, the incidence levels of the prior 4 weeks and prior 5 weeks will be used as predictors in the 4-week ahead prediction.

In addition to the fixed training window approach, we also trained the XGboost algorithm with an extending window. In the fixed window approach, we train the XGBoost model only once with the fixed training set and then predict the test set. Since the data were collected on a weekly basis, we are able to update our model by including the new coming data and retraining it to see if model performance can be improved. Thus we train the model with an extended training set for each week, which is called extending window approach in this paper. For example, if we are going to predict the week *i* (*i* > 2) of the year 2015, our training set used data from 2010 until 2014 for all weeks and data from 2015 for weeks 1 to week (*i* − 1). Our test data were data in 2015 week *i*. The baseline models were trained with the fixed window approach only. Weeks with missing values for predictors were removed from both the training set and the test set.

XGBoost provides a large number of hyperparameters to help achieve optimal performance. They are mainly divided into three types, general parameters, booster parameters, and learning task parameters [42]. Booster parameters are closely related to the performance of the model, which is the focus of hyperparameter tuning. Grid search was used to final optimal values for hyperparameters max_depth, min_child_weight, subsample, colsample_bytree, learning rate and gamma. Grid search can be challenging and time-consuming due to the many parameters to optimize, even with XGBoost’s rapid convergence. Our gird search for hyperparameters was carried out as below:

1) Firstly, find the optimal gamma and eta (learning rate) at the same time since they have an impact on the performance of the model. The values searched for gamma are 0.1, 0.2, 0.5, 1, 1.5, 2, and 10, while those for eta are 0.01, 0.02, 0.03, 0.06, 0.1, 0.2, and 0.3. We ran all possible combinations of these two hyperparameter values to tune, and the one with the best performance was selected as the optimal value for gamma and eta, respectively.

2) With the optimal values of gamma and eta obtained in the previous step, a grid search was conducted for max_depth, and min_child_weight range from 0.1 to 1.

3) Made a grid-search over subsample and colsample_bytree simultaneously range from 0.1 to 1.

K-fold cross-validation is used during the tuning process to assess the model’s performance with different combinations of hyperparameter values. Since there is a dependency between weekly ILI incidence and future values that cannot be used to forecast past values, the traditional K-fold method that randomly splits the training set into K folds is not applicable to our data. Instead, we used cross-validation on an extending basis (S1 Fig). We selected data from 2010 to 2014 as the overall training set for the cross-validation, but during the process of rolling cross-validation, we started with data from 2010 as the first fold of the training set and checked the accuracy of prediction for the data from 2011. Data from 2010 and 2011 then formed the second fold of the training set, and data from 2012 became the test for accuracy check. The accuracy metric used in the tuning process is macro-averaged mean absolute error (mMAE) which will be defined in detail later. This process was repeated until 2014 became the last test set. The final optimal values of the hyperparameter were given in S2 Table.

### Accuracy metrics

We used two of the most commonly used metrics in ordinal classification problems [43], macro-averaged mean absolute error (mMAE) and mean zero-one error (MZE)) to evaluate the model performance. Both metrics are defined as negatively oriented penalties that we aim to minimize: the lower the score, the better the forecast is considered.

#### Macro-averaged mean absolute error

Macro-averaged mean absolute error (mMAE) measure is adapted from the traditional class-based metric mean absolute error (MAE) which assesses the average deviation of the predicted class from the actual class, and it is defined as

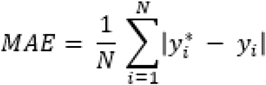

where *y*_*i*_ and 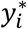 denotes the true class and the predicted class respectively. However, MAE averages effectiveness across individual observations, which does not reflect the imbalanced distribution of classes in our datasets if many observations are in the lowest class and very few are in higher classes. Instead, we used macro-averaged mean absolute error (mMAE) as one of our metrics to assess the performance of our models in terms of predicting ordered outcomes. mMAE is obtained by first computing the MAE on a per-class basis and then averaging the results across the classes so that mMAE is insensitive to class imbalance. Let *y*_*i*_ be the true class and 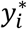 be the predicted class of *i*-th week in the test set. Let *N*_*k*_ be the number of true cases with the class *k* where *k* ∈ {1, 2, …, 9, 10}. There are 10 classes in our classification problem, i.e., 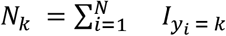 and 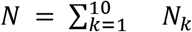 *N*_*k*_. The Macro-averaged mean absolute error is calculated with the following formula:

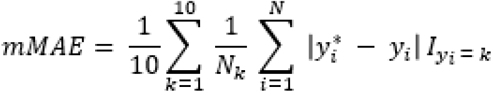

where

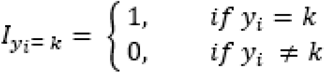

#### Mean zero-one error

Our second metric is the mean zero-one error (MZE) which is more frequently known as the error rate of classifiers and is, essentially, the proportion of time that a correct prediction is made

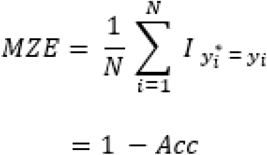

Where

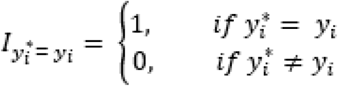

*y*_*i*_ and 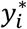 denotes the true class and the predicted class respectively. *Acc* is the accuracy of the model, i.e., the number of correctly predicted categories divided by the total number of predictions. Therefore, MZE ranges between 0 and 1, and the lower the MZE, the higher accuracy indicates better predictive performance.

## Results

Country-level forecasts were performed using ILI data from 32 countries, located in northern temperate regions (S2 Fig). Due to the incompleteness of data, countries located outside of temperate regions were excluded from the analysis. The countries in the northern hemisphere’s temperate regions demonstrated a similar trend in influenza activity, with ILI incidence typically rising at the end of the year and reaching a peak level at the beginning of the following year (Fig 1B). During the 2010 - 2017 surveillance, after excluding weeks with missing values, the data availability ranged between 229 and 417 weeks, with each country contributing an average of 337 weeks of data (median = 391 weeks). Of these weeks, over 60% fell into level 1, while 0.76% of the total weeks fell into levels 9 and 10 (Fig 1A).

**Fig 1.**
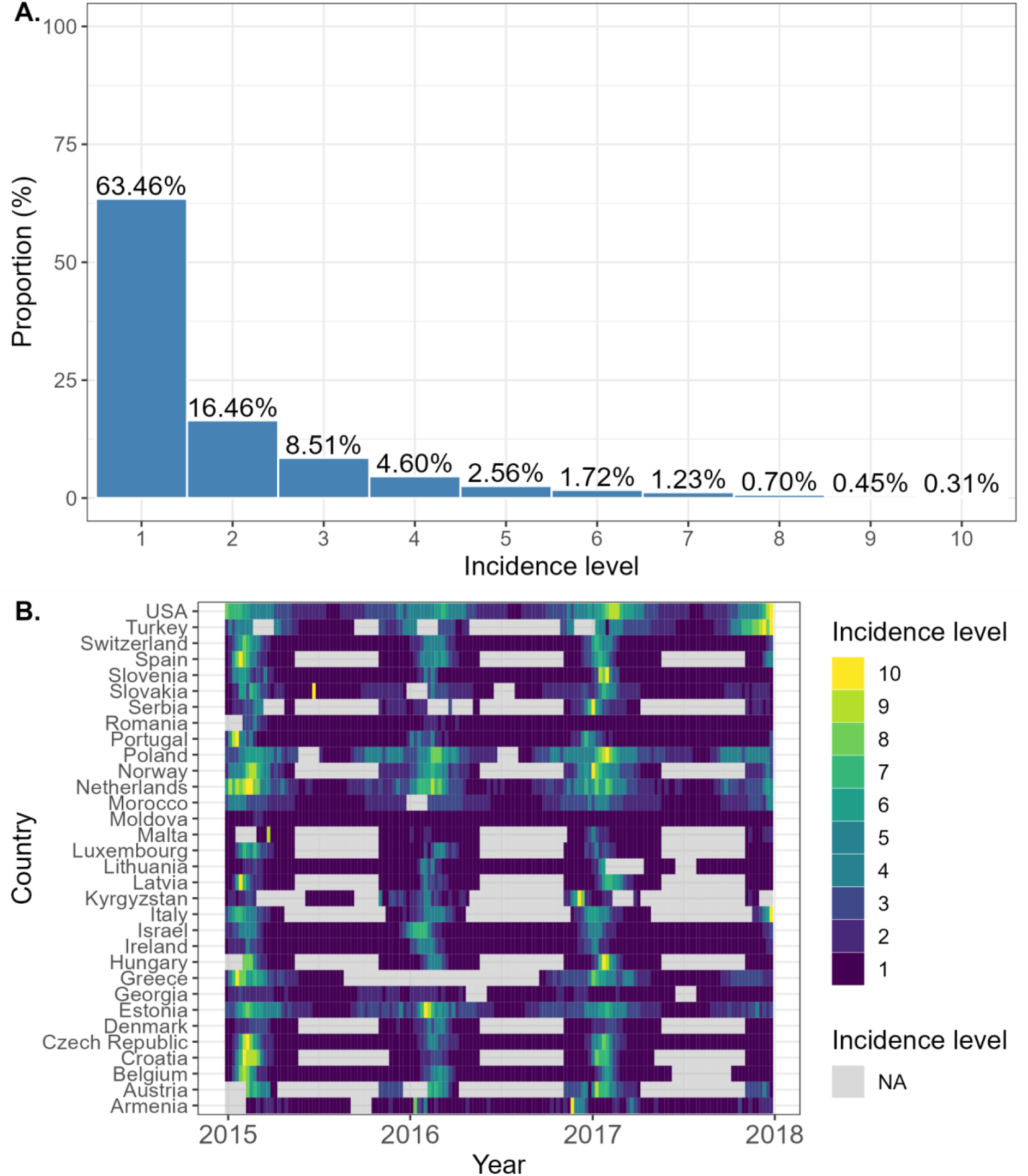
Distribution of categorical incidence levels for weekly influenza-like illness for countries included in the study. **A**. overall frequency of every level appearing in 32 countries. **B**. The heat map shows of incidence level in each country by time. Grey-shaded areas indicate that no available were data for those weeks.

### Predictive performance

Overall, the XGBoost model with an extended window generated fewer errors and demonstrated more stable predictive performance than the XGBoost model with a fixed training window or the baseline models. The short-term prediction errors of different models are summarized in Table 1, by averaging the mMAE is averaged across countries, and presented for each prediction horizon (1-week ahead, 2-week ahead, etc.). We separately compare the mMAEs of three test periods (2015 to 2017). From Table 1, for these three test periods, we show that prediction accuracy measured by mMAEs decreases with the increasing forecast length (i.e. one to four-week in advance), but both XGBoost models (with extending window and fixed window) uniformly outperform the two baseline models (Fig 2, Table 1). In particular, the mMAEs of the XGBoost model with an extended window remain below 1, on average across all countries, even as the forecast length increases, indicating that, on average, the predicted classes are either correct or only one class away from the true class. This XGBoost model has the lowest average MZEs for all prediction horizons compared to the XGBoost model with a fixed window and baseline models (S3 Fig, S3 Table).

**Table 1.**
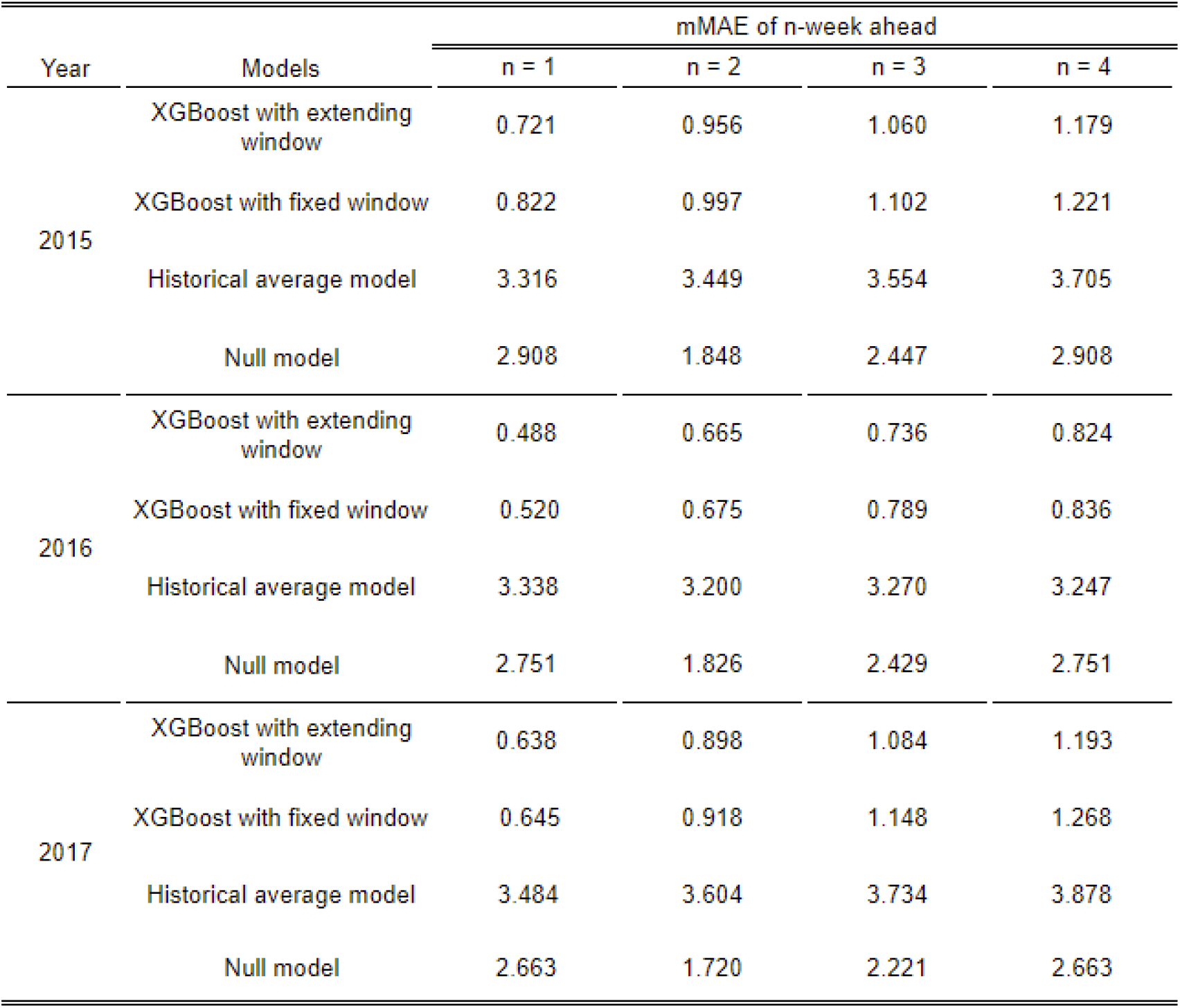
Overall macro-averaged mean absolute error (mMAE) of XGboost compared with baseline models. Overall mMAEs of 1- to 4-week ahead forecasts for each model are calculated as the average of 32 countries’ mMAEs by year.

| Year | Models | mMAE of n-week ahead |  |  |  |
| --- | --- | --- | --- | --- | --- |
|  |  | n = 1 | n = 2 | n = 3 | n = 4 |
| 2015 | XGBoost with extending window | 0.721 | 0.956 | 1.060 | 1.179 |
|  | XGBoost with fixed window | 0.822 | 0.997 | 1.102 | 1.221 |
|  | Historical average model | 3.316 | 3.449 | 3.554 | 3.705 |
|  | Null model | 2.908 | 1.848 | 2.447 | 2.908 |
| 2016 | XGBoost with extending window | 0.488 | 0.665 | 0.736 | 0.824 |
|  | XGBoost with fixed window | 0.520 | 0.675 | 0.789 | 0.836 |
|  | Historical average model | 3.338 | 3.200 | 3.270 | 3.247 |
|  | Null model | 2.751 | 1.826 | 2.429 | 2.751 |
| 2017 | XGBoost with extending window | 0.638 | 0.898 | 1.084 | 1.193 |
|  | XGBoost with fixed window | 0.645 | 0.918 | 1.148 | 1.268 |
|  | Historical average model | 3.484 | 3.604 | 3.734 | 3.878 |
|  | Null model | 2.663 | 1.720 | 2.221 | 2.663 |

**Fig 2.**
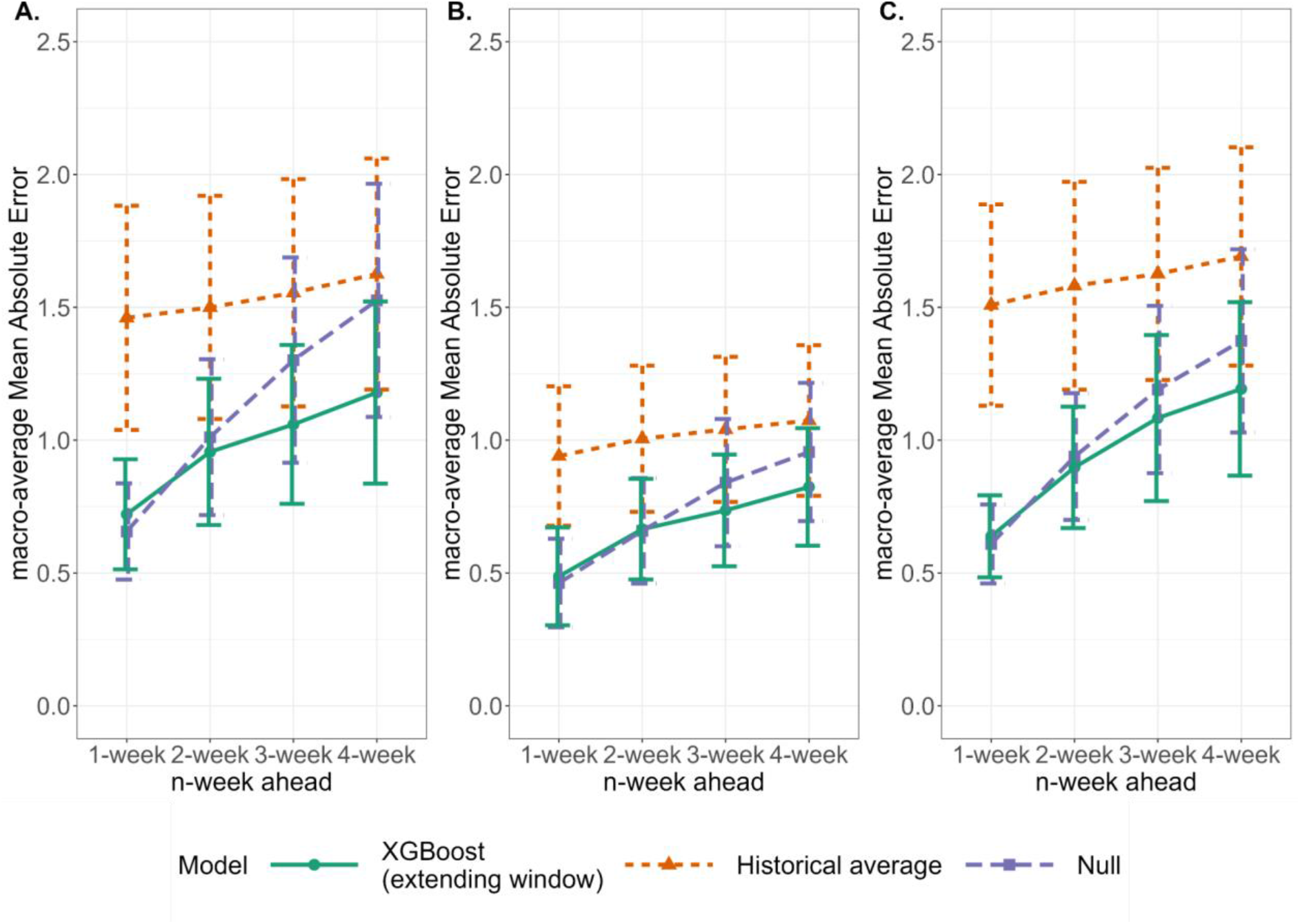
Differences in macro-averaged mean absolute error (mMAE) by model. Comparison of mMAE for XGBoost (with an extended window) and baseline models while forecasting 1 to 4 weeks ahead. **A**. 2015, **B**. 2016, **C**. 2017.

The mMAEs for the extending window approach were consistently lower compared to the XGBoost model with a fixed window (Fig 3, Table 1). The mMAEs for the fixed window approach only remained below 1 for 1-to 4-week forecasts in 2016, but exceeded 1 for 3- and 4-week ahead forecasts in 2015 and 2017. Although the two XGBoost models had similar MZEs, the extending window approach still resulted in a lower MZE for each prediction horizon (S4 Fig, S3 Table). The better performance of the extending window approach can be attributed to its continuous addition of the predicted weeks’ observations to the training set, allowing the model to learn more data, resulting in smaller prediction errors.

**Fig 3.**
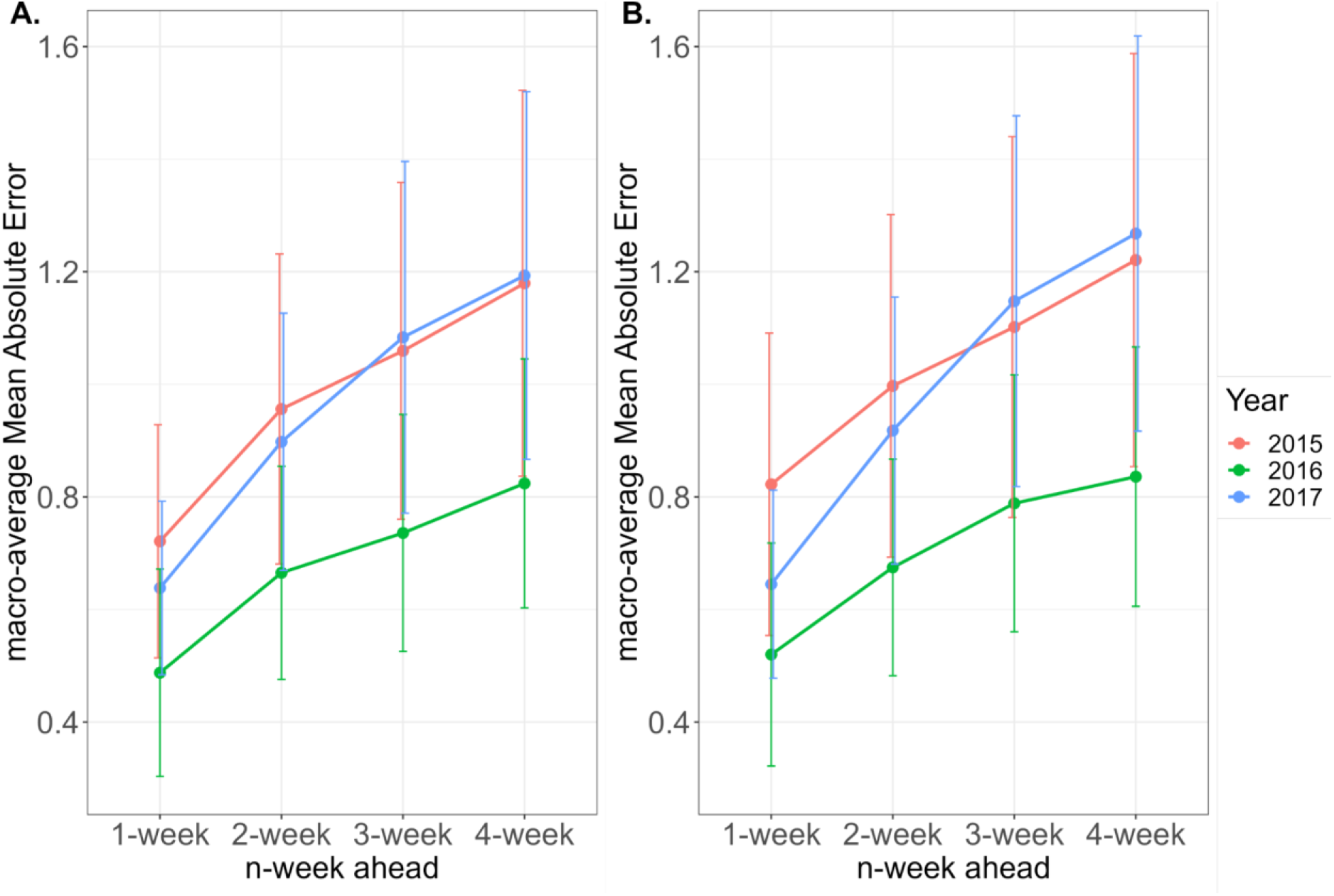
Macro-averaged mean absolute error (mMAE) of XGBoost models. Comparison of mMAEs for weekly forecasting ranging from 1-to 4-week ahead for the years 2015 to 2017. **A**. The XGBoost model with an extended window approach. **B**. The XGBoost model with a fixed window approach.

There is substantial variation in the prediction accuracy, as indicated by mMAE, among the countries (Fig 4, S4 – S6 Table). For instance, Moldova, had remarkably low mMAEs from 2015 to 2017, even achieving 0 in 2016 and 2017. Conversely, certain countries, such as Hungary and Norway, consistently exhibit much higher mMAEs compared to other countries. Similar results were found for MZEs (S5 Fig, S7 – S9 Table). The performance of XGBoost on the test set can be affected mainly by the quality of training data. By checking the missing data for countries, we discover that countries with high mMAEs often have more incomplete data than better-performing countries, implying that they have a smaller sample size of training sets, failing to capture complex epidemic patterns of influenza (Fig 2B).

**Fig 4.**
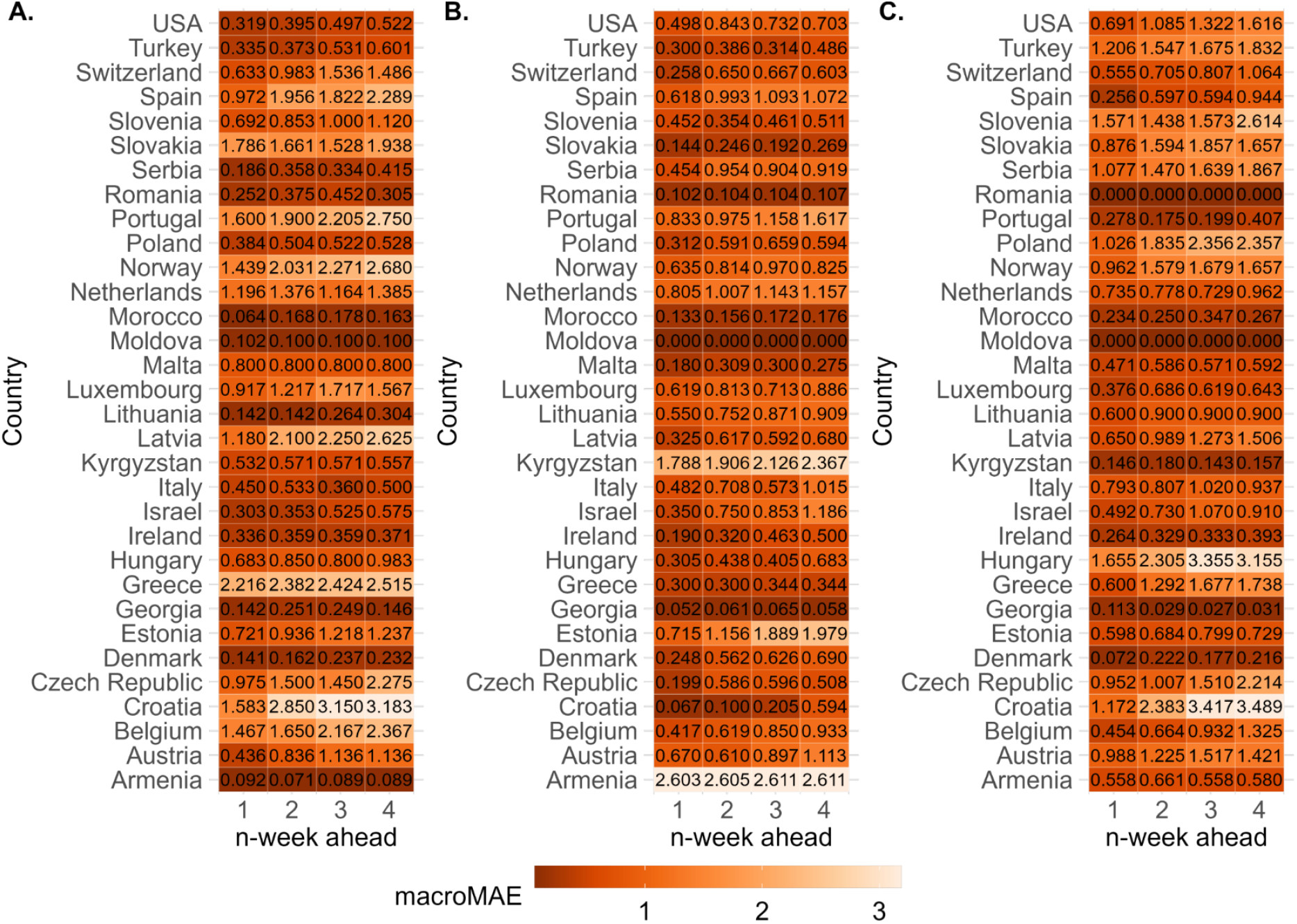
Macro-averaged mean absolute error (mMAE) for 32 countries. mMAEs of predictions by the XGBoost model with an extended window for the year **A**. 2015. **B**. 2016. and **C**. 2017.

We investigated the influenza distribution and forecasts over a 3-year period in four countries selected based on their mMAEs and data completeness and found different qualitative drivers of accuracy. The plots in Fig 5 compare the predicted values from four models with actual data for 1 to 4-week-ahead forecasts in Moldova, Switzerland, Estonia and Hungary separately. Moldova had the lowest mMAE, but its influenza activity barely fluctuated with 97.5% (390 out of 400 weeks) of the weeks remaining at level 1. Conversely, Switzerland and Estonia had 25.3% (82 out of 406 weeks) and 72.5% (290 out of 400 weeks) of the weeks, respectively, above the baseline level. All four models successfully predict peak influenza season for 1-week and 2-week ahead forecasts in 2015 and 2016 in

**Fig 5.**
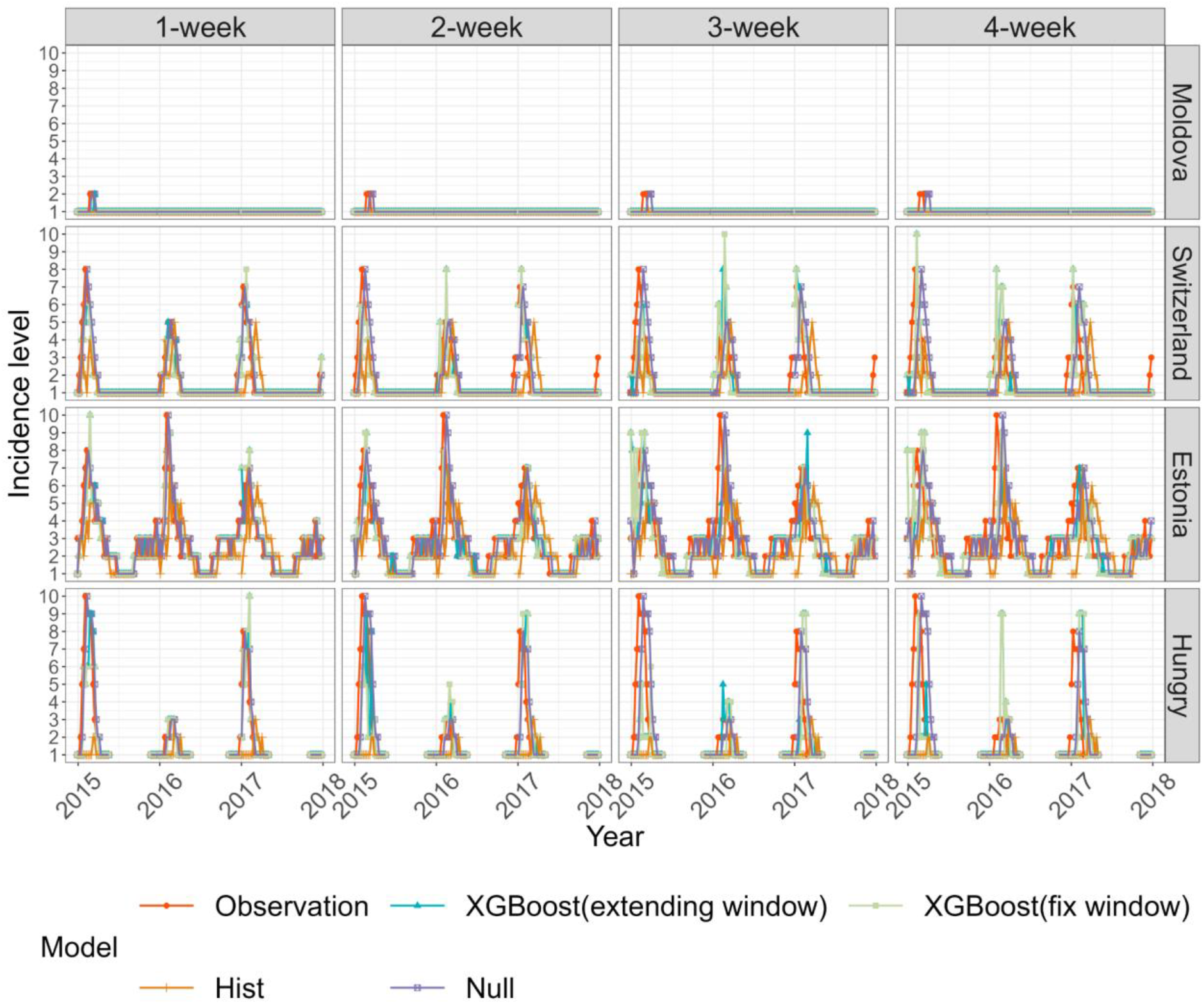
Comparison of Actual and predicted incidence by country. Countries Moldova, Switzerland, Estonia and Hungary are selected. Models include the two XGBoost models, the historical average model and the null model while forecasting 1-to 4-week ahead for the test period year 2015 to 2017.

Switzerland Estonia, but the historical average model failed to identify peaks for 3-week and 4-week ahead forecasts. The other three models could identify peaks up to 4-week ahead of forecasts, with the XGboost with extending approach being the most accurate. The deviation between actual observation and prediction is typically high when there is a sudden increase or decrease in flu incidence levels. In Hungary, due to the limited sample size of the training set and the complexities of influenza activity, a large deviation between actual observation and prediction even for the 1-week forward forecast. The prediction of XGboost with an extending window provided the closest prediction to the observed peak (Fig 6).

**Fig 6.**
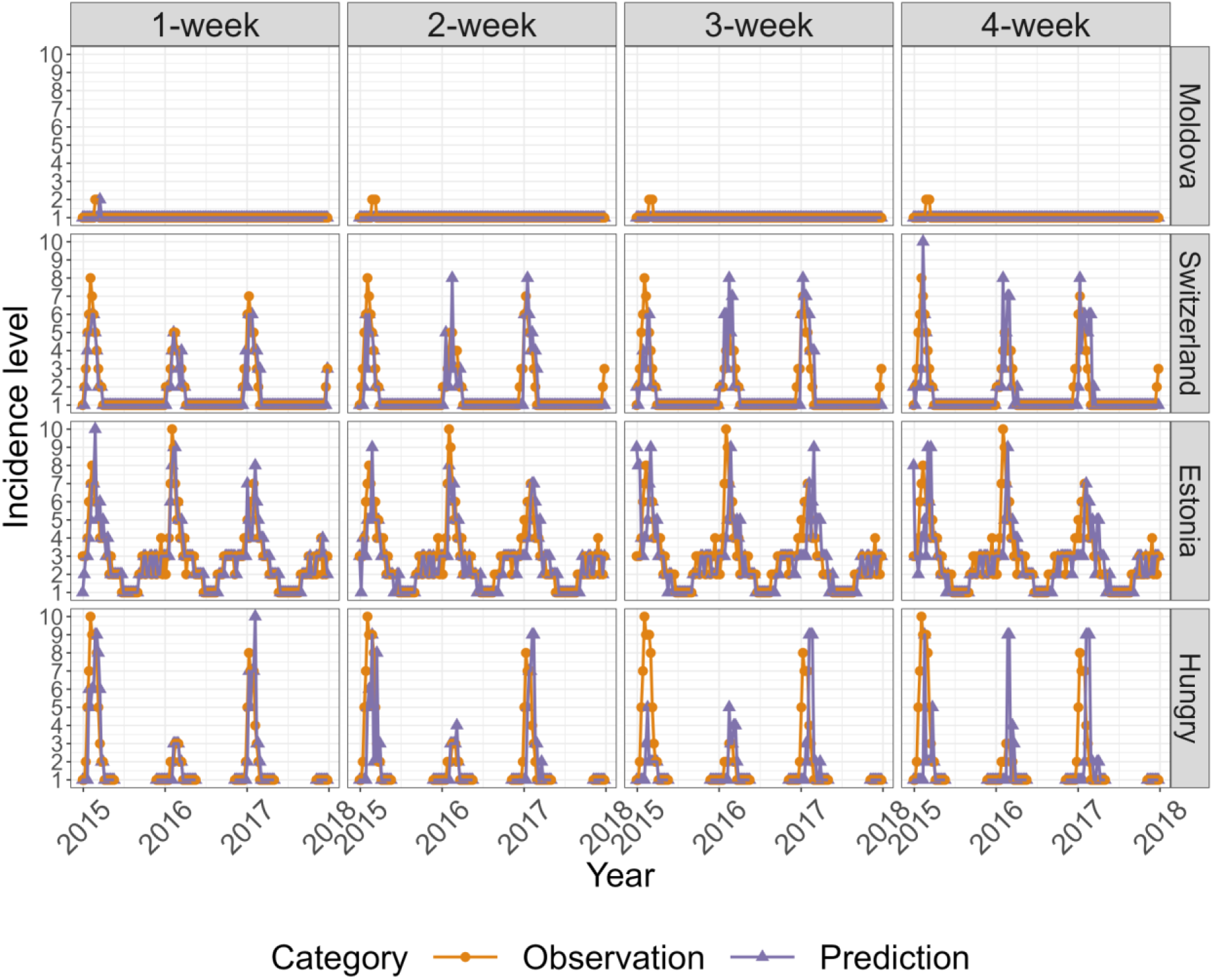
Comparison of actual and predicted incidence. Predicted incidences are generated by the XGBoost model with an extended window for Moldova, Switzerland, Estonia and Hungry; for forecasts 1 to 4 weeks ahead for the test period year 2015 to 2017.

## Discussion

In this work, we have proposed a potential machine learning model for influenza forecasting using a 10-unit ordinal variable to describe the case incidence. We evaluated two XGBoost machine learning models, one with a fixed window training set and the other with an extended window training set, and compared them to baseline models. We measured their accuracy using the mean zero-one error (MZE) and macro-averaged mean absolute error (mMAE). Our results showed that both XGBoost models outperformed the baseline models, with the extended window XGBoost model consistently achieving the highest accuracy.

This framework is novel because we defined the outcome on an ordinal scale, which is often how influenza incidence is communicated[32]. However, our results are not directly comparable to other forecasting studies, and the XGBoost model has not been validated against other data sources. This highlights the need to test its applicability in real-world public health practice.

The use of a categorical outcome enabled us to apply machine learning algorithms. Although the XGBoost has advantages, it has not been tested against other data sources. Machine learning algorithms’ performance can be limited by the quality of the training and testing data, and inconsistencies between different databases may reduce the generalisability of the XGBoost prediction model [44]. Further testing is needed to determine the applicability of the in real-world public health practice.

Our baseline models provide a basic framework for assessing the potential of machine learning forecasts. The baseline models are intended to reflect the most basic of implicit forecast assumptions: they assume that incidence will remain unchanged and revert to its historical average. Our methods offer improvements over these assumptions, for example, the 4-week forecast horizon XGBoost model with an extended window being more accurate than assuming the incidence would follow the historical average and more accurate than the default 1-week ahead model with no change assumption.

This work has several limitations. One of the challenges in working with infectious disease datasets is the limited period of data available for model estimation and evaluation, compared to other datasets used in machine learning. For each country in the study, only 5 years of data were used for model training, leaving only 3 years for testing. Cross-validation was used to find the optimal hyperparameters instead of using an independent validation set. Additionally, countries with fewer weeks of data in the training set tend to have higher prediction errors, which suggests that accuracy will improve as more data becomes available. Furthermore, the results should be interpreted with caution, given the disruption to influenza transmission during the COVID-19 pandemic, as patterns may change as the transmission is re-established.

Our forecasts showed reduced accuracy in predicting turning points of the epidemic (Fig 7). When there is a sudden increase in incidence levels, they are often underestimated. On the other hand, a sudden decrease in incidence levels results in them being easily overestimated. This is especially important in a public health context where accurately predicting rare cases of much higher than usual incidence levels or earlier or later ends of the epidemic is crucial. Thus, improving the prediction stability at the extreme points of season patterns remains a priority in ongoing forecasting work.

**Fig 7.**
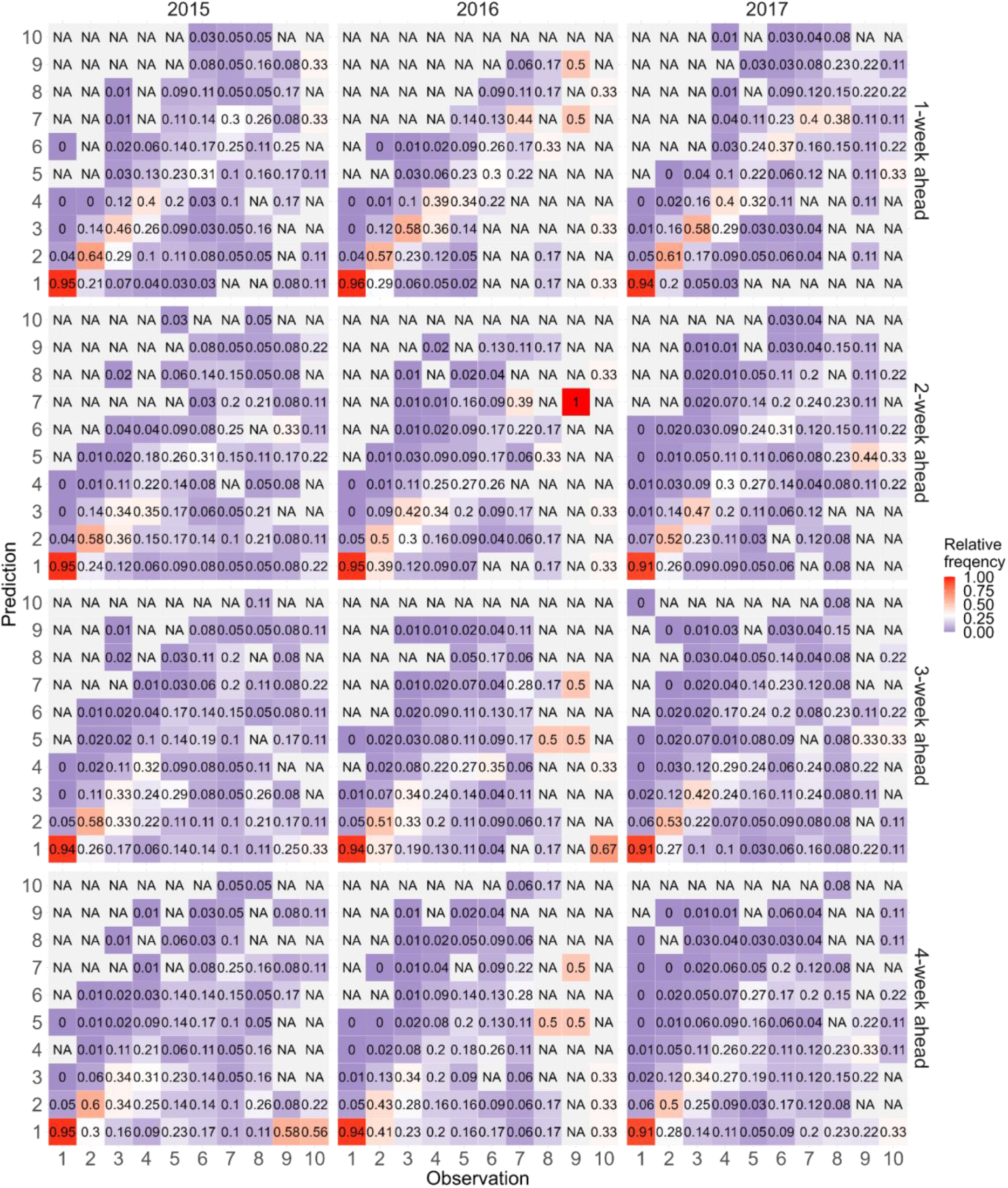
Distribution of predictions against each decile of observation for the XGBoost model with an extended window. Rows of heat maps are ordered from 1 week ahead (top) to 4 weeks ahead (bottom). Columns of heat maps are ordered from 2015 (left) to 2017 (right). Values on the diagonal are the accuracy of forecasts, while other values represent the frequency at each level that was incorrectly predicted to be the other level.

## Conclusion

Forecasting influenza as an ordinal outcome is a feasible task for machine learning. The widely used XGBoost model, even with a limited set of features, provides significantly more accurate predictions than the standard baseline models. With datasets that have longer history and comprehensive spatial coverage, it is possible to achieve more accurate forecasts. Similar to other epidemiological models, the framework can easily be expanded to include population serology and population mobility [45] or other relevant features as more data become available [46].

## Data Availability

All data produced are available online at https://zenodo.org/badge/latestdoi/597533400

## Supporting information

**S1 Table. Time ranges of training and test sets**.

(XLSX)

**S2 Table. Summary of optimal values of hyperparameters for XGBoost models**.

(XLSX)

**S3 Table. Overall mean-zero error (MZE) of XGboost compared with baseline models**. Overall MZEs of 1-to 4-week ahead forecasts for each model are calculated as the average of 32 countries’ MZEs by year.

(XLSX)

**S4 Table. Macro-average mean absolute error (mMAE) for the prediction by country in 2015**.

(XLSX)

**S5 Table. Macro-average mean absolute error (mMAE) for the prediction by country in 2016**.

(XLSX)

**S6 Table. Macro-average mean absolute error (mMAE) for the prediction by country in 2017**.

(XLSX)

**S7 Table. Mean-zero error (MZE) for the prediction by country in 2015**.

(XLSX)

**S8 Table. Mean-zero error (MZE) for the prediction by country in 2016**.

(XLSX)

**S9 Table. Mean-zero error (MZE) for the prediction by country in 2017**.

(XLSX)

**S1 Fig.**
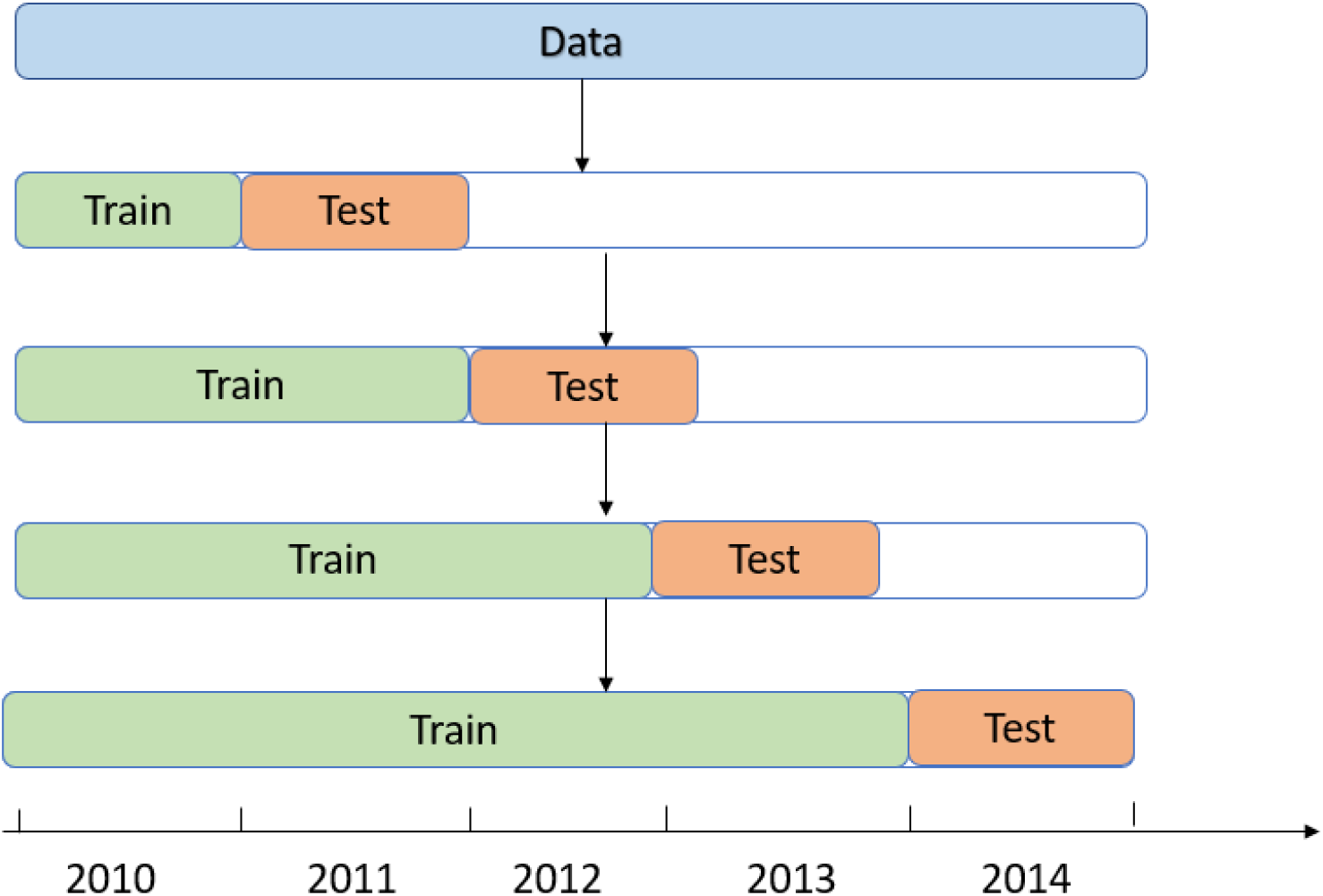
Time series split cross-validation.

**S2 Fig.**
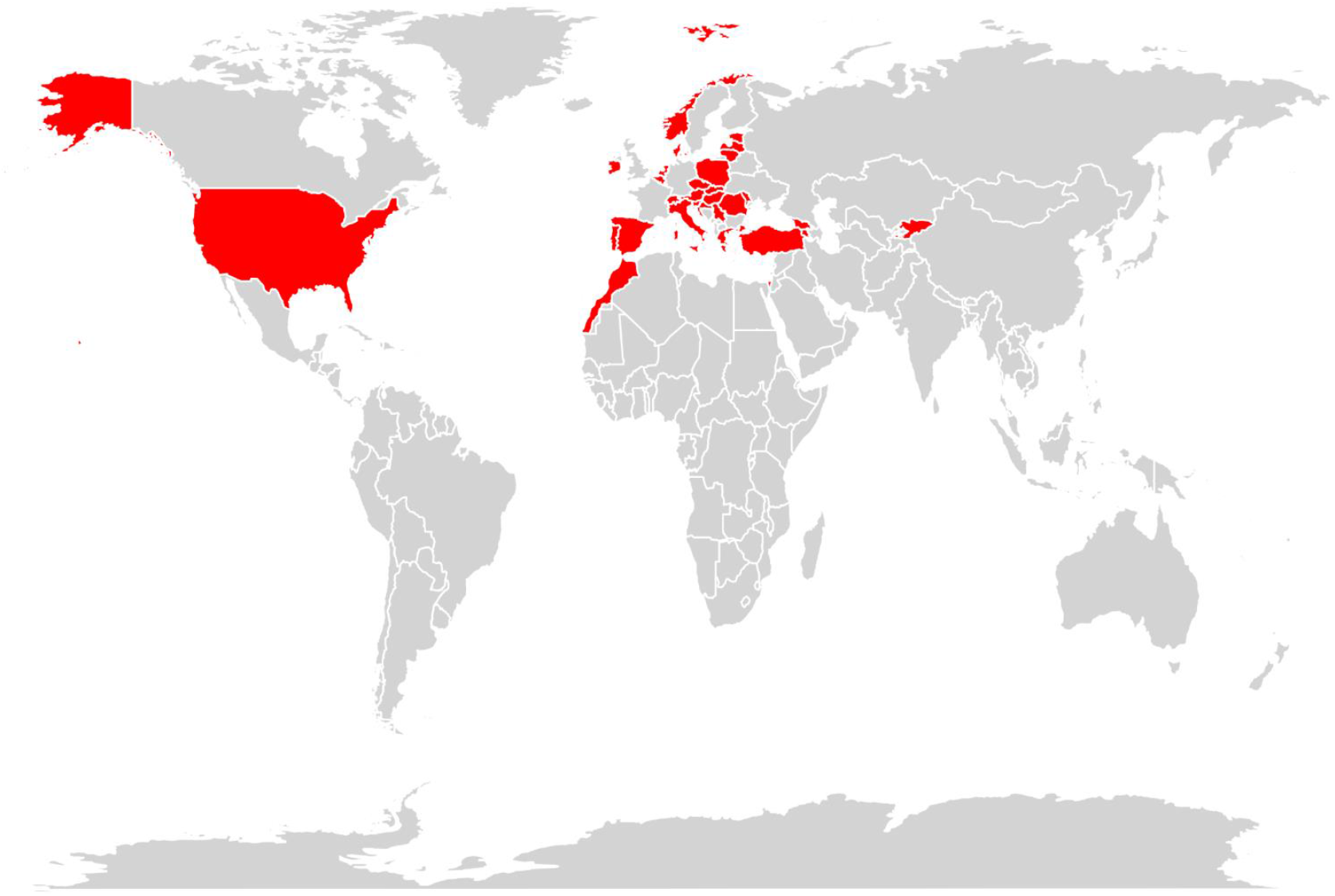
World map. 32 Countries included in this study are marked in red. Samples are mainly from North America and Europe.

**S3 Fig.**
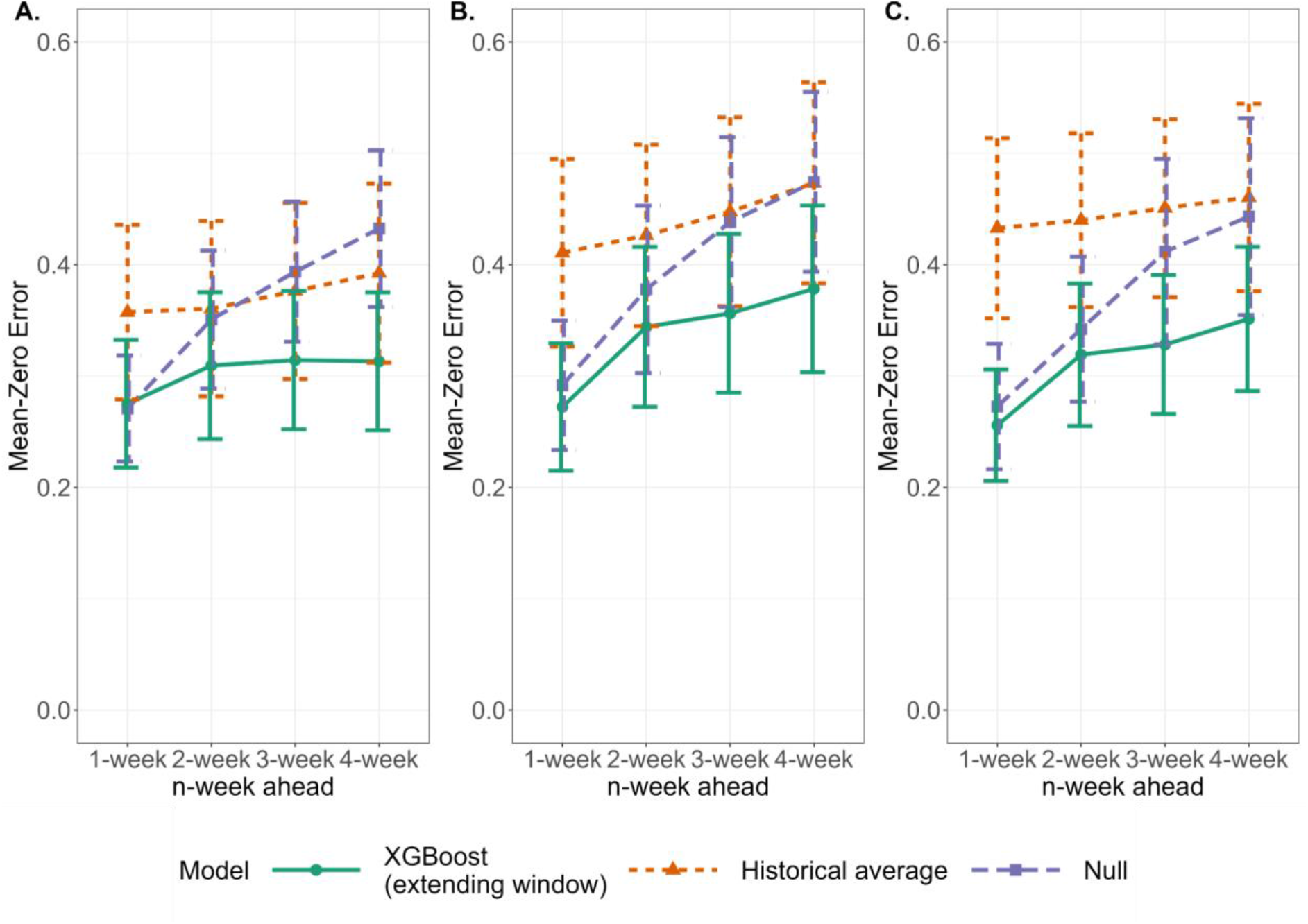
Differences in mean-zero error (MZE) by model. Comparison of mean-zero error (MZE) for XGBoost (with an extended window approach) and baseline models while forecasting 1 to 4 weeks ahead for the test period year 2015 to 2017.

**S4 Fig.**
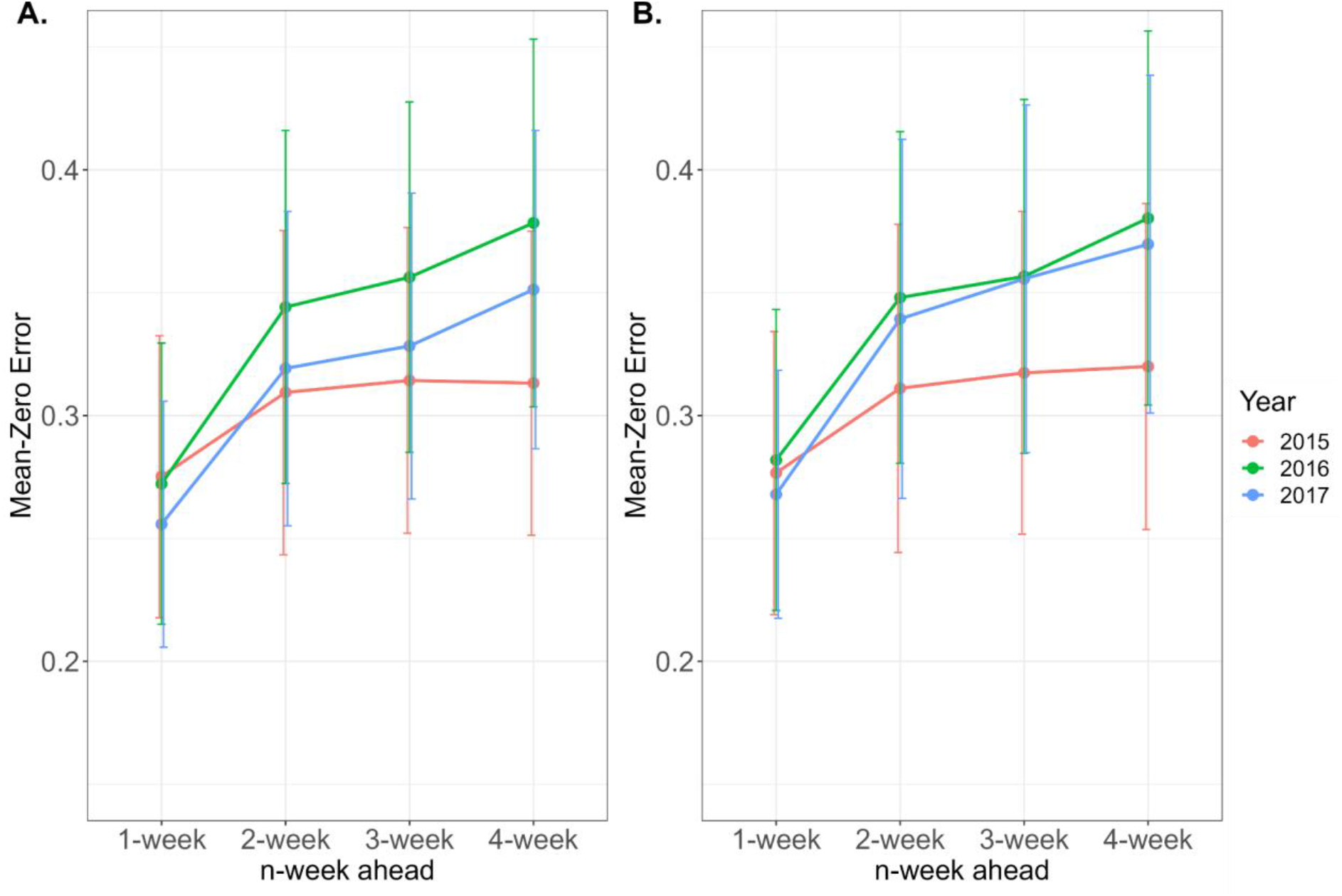
Mean-zero error (MZE) of XGBoost models. Comparison of MZEs for weekly forecasting ranging from 1-to 4-week ahead in 2015 to 2017. **A**. The XGBoost model with an extended approach. **B**. The XGBoost model with a fixed window approach.

**S5 Fig.**
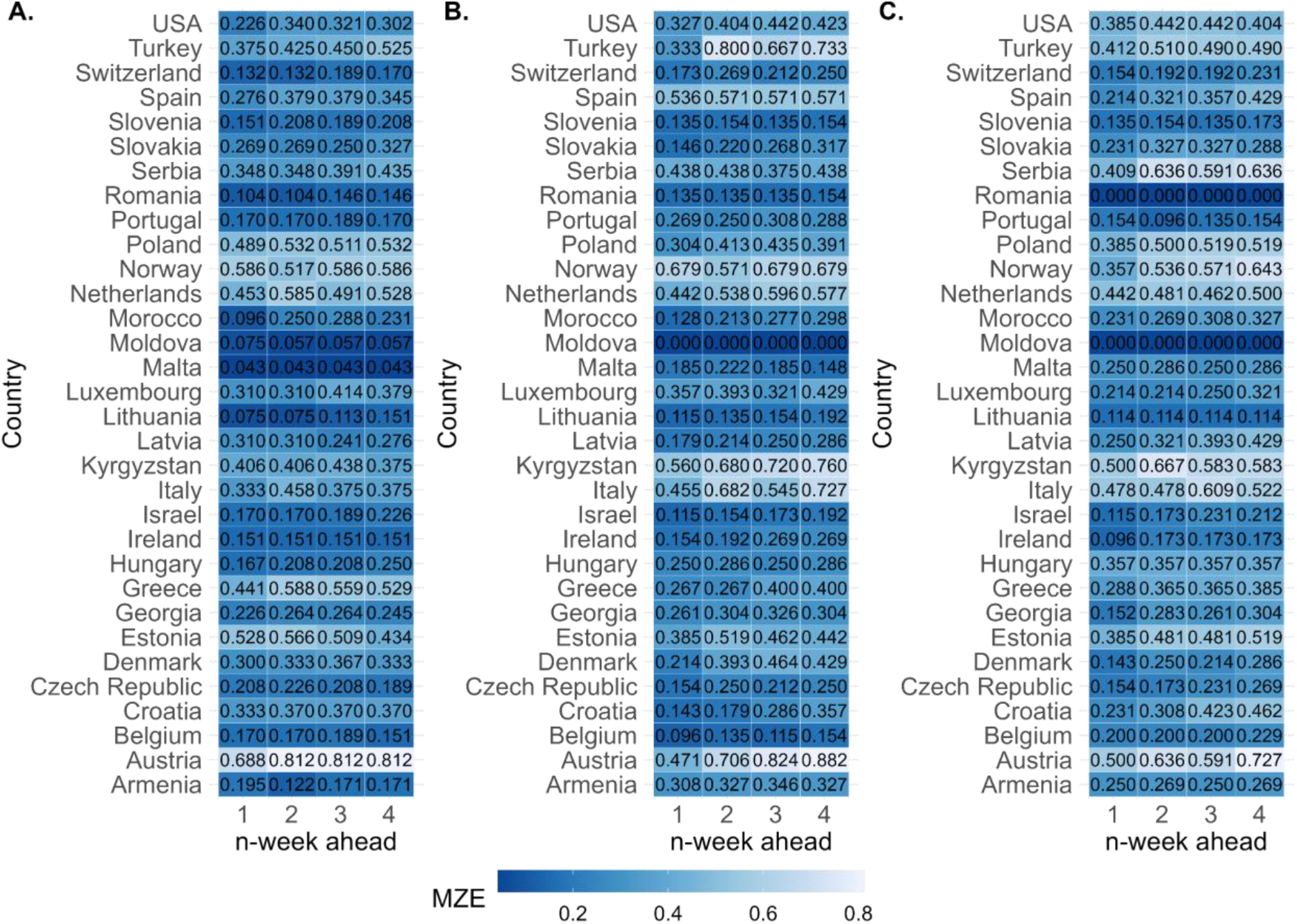
Mean-zero error (MZE) for 32 countries. MZEs of predictions by the XGBoost model with an extended window for the year **A**. 2015. **B**. 2016. and **C**. 2017.

## Acknowledgements

The authors acknowledge funding from the MRC Centre for Global Infectious Disease Analysis (MR/R015600/1), jointly funded by the UK Medical Research Council (MRC) and the UK Foreign, Commonwealth & Development Office (FCDO), under the MRC/FCDO Concordat agreement, and also part of the EDCTP2 programme supported by the European Union. S.R. acknowledges the support from Wellcome Trust Investigator Award (UK, 200861/Z/16/Z). KOK acknowledges funding from HMRF (INF-CUHK-1).

## Author Contributions

**Conceptualization:** Haowei Wang, Steven Riley

**Data curation:** Haowei Wang

**Formal analysis:** Haowei Wang

**Funding acquisition**

**Methodology:** Haowei Wang

**Software:** Haowei Wang

**Supervision:** Steven Riley

**Validation:** Haowei Wang

**Visualization:** Haowei Wang

**Writing – original draft:** Haowei Wang

**Writing – review & editing:** Haowei Wang, Steven Riley, Kin On Kwok

## Notes

### Competing Interest Statement

The authors have declared no competing interest.

